# SARS-CoV-2 wastewater variant surveillance: pandemic response leveraging FDA’s GenomeTrakr network

**DOI:** 10.1101/2024.01.10.24301101

**Authors:** Ruth E. Timme, Jacquelina Woods, Jessica L Jones, Kevin R Calci, Rachel Rodriguez, Candace Barnes, Elizabeth Leard, Mark Craven, Haifeng Chen, Cameron Boerner, Christopher Grim, Amanda M. Windsor, Padmini Ramachandran, Tim Muruvanda, Hugh Rand, Bereket Tesfaldet, Jasmine Amirzadegan, Tunc Kayikcioglu, Tamara Walsky, Marc Allard, Maria Balkey, C. Hope Bias, Eric Brown, Kathryn Judy, Tina Pfefer, Sandra M Tallent, Maria Hoffmann, James Pettengill, the GenomeTrakr Laboratory consortium

**Affiliations:** Office of Regulatory Science, Center for Food Safety and Applied Nutrition, US Food and Drug Administration, College Park, MD, USA; Gulf Coast Seafood Laboratory, Center for Food Safety and Applied Nutrition, US Food and Drug Administration, Dauphin Island, AL, USA; Office of Applied Research SA, Center for Food Safety and Applied Nutrition, US Food and Drug Administration, Laurel, MD, USA; Office of Analytics and Outreach, Center for Food Safety and Applied Nutrition, US Food and Drug Administration, College Park, MD, USA; Oak Ridge Institute for Science and Education, U.S. Department of Energy, Oak Ridge, TN, USA; Joint Institute for Food Safety and Applied Nutrition (JIFSAN), University of Maryland – College Park, College Park, MD, USA

## Abstract

Wastewater surveillance has emerged as a crucial public health tool for population-level pathogen surveillance. Supported by funding from the American Rescue Plan Act of 2021, the FDA’s genomic epidemiology program, GenomeTrakr, was leveraged to sequence SARS-CoV-2 from wastewater sites across the United States. This initiative required the evaluation, optimization, development, and publication of new methods and analytical tools spanning sample collection through variant analyses. Version-controlled protocols for each step of the process were developed and published on protocols.io. A custom data analysis tool and a publicly accessible dashboard were built to facilitate real-time visualization of the collected data, focusing on the relative abundance of SARS-CoV-2 variants and sub-lineages across different samples and sites throughout the project. From September 2021 through June 2023, a total of 3,389 wastewater samples were collected, with 2,517 undergoing sequencing and submission to NCBI under the umbrella BioProject, PRJNA757291. Sequence data were released with explicit quality control (QC) tags on all sequence records, communicating our confidence in the quality of data. Variant analysis revealed wide circulation of Delta in the fall of 2021 and captured the sweep of Omicron and subsequent diversification of this lineage through the end of the sampling period. This project successfully achieved two important goals for the FDA’s GenomeTrakr program: first, contributing timely genomic data for the SARS-CoV-2 pandemic response, and second, establishing both capacity and best practices for culture-independent, population-level environmental surveillance for other pathogens of interest to the FDA.

**IMPORTANCE:** This manuscript serves two primary objectives. Firstly, it summarizes the genomic and contextual data collected during a Covid-19 pandemic response project, which utilized the FDA’s laboratory network, traditionally employed for sequencing foodborne pathogens, for sequencing SARS-CoV-2 from wastewater samples. Secondly, it outlines best practices for gathering and organizing population-level Next Generation Sequencing (NGS) data collected for culture-free, surveillance of pathogens sourced from environmental samples.

## INTRODUCTION

All viruses, including severe acute respiratory syndrome coronavirus 2 (SARS-CoV-2) evolve over time, accumulating random mutations within their genomes that result in new variants and lineages. Although tracking the early spread of SARS-CoV-2 was primarily done through PCR tests, sequencing the entire genome facilitates the identification and tracking of new mutations and lineages. This is especially important when those mutations alter clinical characteristics, such as replicating faster than others, causing different symptoms or severity of disease, or eluding vaccines or therapeutic treatments. In early 2021 the first “variants of concern” started to emerge from SARS-CoV-2 (1), e.g., Alpha (B.1.1.7), Beta (B.1.351), and (Gamma) P.1. Suddenly, merely testing for the presence of the virus wasn’t sufficient to track the pandemic. The full genome sequence became necessary to identify new mutations, emerging variants, and sub-lineages.

FDA’s GenomeTrakr Program (2), a pathogen genomic surveillance network led by the FDA Center for Food Safety and Applied Nutrition (CFSAN), has been collaborating with other US Government and State public health agencies (3) to use whole genome sequence data to ensure food safety and assist with epidemiological investigations of foodborne pathogens since 2012. This laboratory network comprises 31 federal and state public health laboratories, each equipped with the instrumentation and trained personnel required for pathogen sequencing and data submission to the NIH’s National Center for Biotechnology Information (NCBI). By design, the network is focused on sequencing pathogens from food samples, food facilities, the farm environment, and adjacent waterways. Resulting genomic data informs regulatory decisions around foodborne disease outbreaks or food production environments. A dedicated funding model supports these activities, which include submitting raw sequence data along with a minimum set of contextual data to the publicly accessible NCBI database in real time (4). This model, while good general practice for a publicly funded pathogen surveillance network, is also an ideal model for the rapid sharing of pathogen genome sequence data during a global pandemic.

Although SARS-CoV-2 is not a virus that causes foodborne illness, several factors contributed to the tapping of GenomeTrakr to leverage its laboratory network for sequencing SARS-CoV-2 genomes and assist efforts of the U.S. Government to better monitor the spread of new SARS-CoV-2 variants and mutations. Funding for this work came from the American Rescue Plan Act of 2021, which included public health funding for pandemic response. Wastewater was chosen as a surveillance tool for multiple reasons. It is optimal for acquiring timely population-level sequence data given its full suite of circulating and emerging mutations, which are valuable for independent validation and verification of FDA approved therapeutics, diagnostics, and vaccines. New lineages of SARS-CoV-2 can be identified in wastewater samples up to a week prior to being detected in health-care seeking individuals from the same population (5–7). Routine wastewater samples also provide a relatively unbiased capture of genomic variation from the entire sewage catchment area, as opposed to clinical samples of a given population providing limited information on circulating variants. These samples may also reveal cryptic lineages not seen in the clinical sequence database (8). Furthermore, site locations can be targeted for monitoring specific types of populations (e.g. food production and agriculture workers). Choosing wastewater sites that captured circulating SARS-CoV-2 among these populations would meet these goals for the FDA and compliment efforts by the Centers for Disease Control and Prevention (CDC) and regional partners, which were initially focused on urban sewer sheds.

Our goal here is to provide an overview of FDA’s efforts to perform timely wastewater surveillance for SARS-CoV-2 by leveraging the existing GenomeTrakr laboratory network, as well as provide some lessons learned in this endeavor. We also give an overview of the sequence data collected to date from our systematic random wastewater sampling, identify which laboratory methods yielded high quality data, and describe our best practices for implementing these methods within a public health setting.

## METHODS

Five major steps were necessary to build capacity for timely SARS-CoV-2 wastewater surveillance by the GenomeTrakr sequencing laboratories: 1) fund GenomeTrakr laboratories recruited for this project; 2) test, optimize, develop, and publish new laboratory methods for sequencing population-level SARS-CoV-2 from wastewater samples; 3) develop and publish data analysis methods that accessed the sequence quality of raw data and predicted proportions of SARS-CoV-2 variants within each sample; 4) develop and publish protocols for timely data submission to NCBI; and 5) create a public dashboard to visualize variant data from those routine data submissions across the network, providing timely data release and data analysis for public health applications.

### Laboratory funding and site selection

GenomeTrakr laboratories are supported by the FDA Laboratory Flexible Funding Model (9). With additional funding provided by the American Rescue Plan Act of 2021, in the spring of 2021, participating labs were invited to apply to participate in this special pandemic response wastewater project. Participating labs were required to select a minimum of two regional wastewater sites for routine sample collection, which involved sampling 1-2 times per week over a period of at least 6 months. The SARS-CoV-2 population in each sample would be sequenced, following RT-qPCR detection, and labs would submit both their sequencing data and a suite of rich contextual data to the NCBI as soon as possible. The selected regional wastewater sites were chosen in an attempt to capture areas within each respective state that had higher populations of food and agriculture workers, assisted by county-level maps generated within FDA’s 21 *FORWARD* (10) data platform.

### Laboratory method development

In 2021, methods for enriching and sequencing SARS-CoV-2 from wastewater samples were in the early stages of being developed, with most laboratories focused on adapting targeted amplification panels used for clinical sequencing (11–13) to wastewater samples and a few also exploring oligo-capture approaches (14). A comprehensive set of standardized procedures for the entire wastewater processing workflow was needed. This workflow included sample collection, detection and quantification of SARS-CoV-2, SARS-CoV-2 sequencing, analysis, data submission, and visualization. Existing methods covering this workflow were tested, optimized, and published within the GenomeTrakr workspace on protocols.io (15). This platform facilitated real-time communication with version control to our laboratories and to the broader community. In total, 16 new protocols were drafted and published including wastewater sample collection (16), concentration and nucleic acid extraction (17–20), SARS-CoV-2 detection by RT-qPCR (21–23), and SARS-CoV-2 targeted amplification and sequencing (24–27). As long as the participating laboratories adhered to the tiled amplicon + short read sequencing approach established for this project, they had the option of adopting our methods and following our protocols or using different methods of their choice.

### Quality Control

At the start of this project in 2021, quality control (QC) checkpoints in the laboratory workflow as well as final QC thresholds for sequence data of SARS-CoV-2 from a mixed population sample had not yet been defined. One project objective was to identify those crucial QC checkpoints within the laboratory workflow and define thresholds for PASS/FAIL at each of these steps that would yield data of sufficient quality to calculate relative abundance of circulating variants within a given sample.

### NCBI data structure

Raw sequence data plus an extensive suite of contextual data describing the wastewater catchment area, site location information, methods for sampling, extracting nucleic acids, and sequencing the target pathogen all need to be structured and standardized so that data could be compared within our study and most importantly, among studies. To ensure our data was FAIR (findable, accessible, interoperable, and reusable) (28), we defined a standard data structure, or “Data Object Model” (DOM) for pathogen-targeted sequence data from environmental sources. To accomplish this, we modified an existing DOM widely used for genomic pathogen surveillance (4, 29). This environmental pathogen DOM is a standard structure, or “open protocol”, providing interoperability across public and private data repositories for population-level pathogen sequence data collected from environmental sources (wastewater, water, soil, air, etc) (Figure 1). This data structure includes a BioProject describing the scope of study (for our study, one BioProject per lab). Linked to the BioProject are a set of BioSamples set at the nucleic acid extraction level. These BioSample records comprise a variety of sample attributes, including the geographic location where the water was collected, specific site information, and sampling/concentration/and nucleic acid methods. Lastly, raw sequence data along with contextual data describing the experimental sequencing methods, filtering, and QC assessment are linked to the BioSample records.

**Figure 1.**
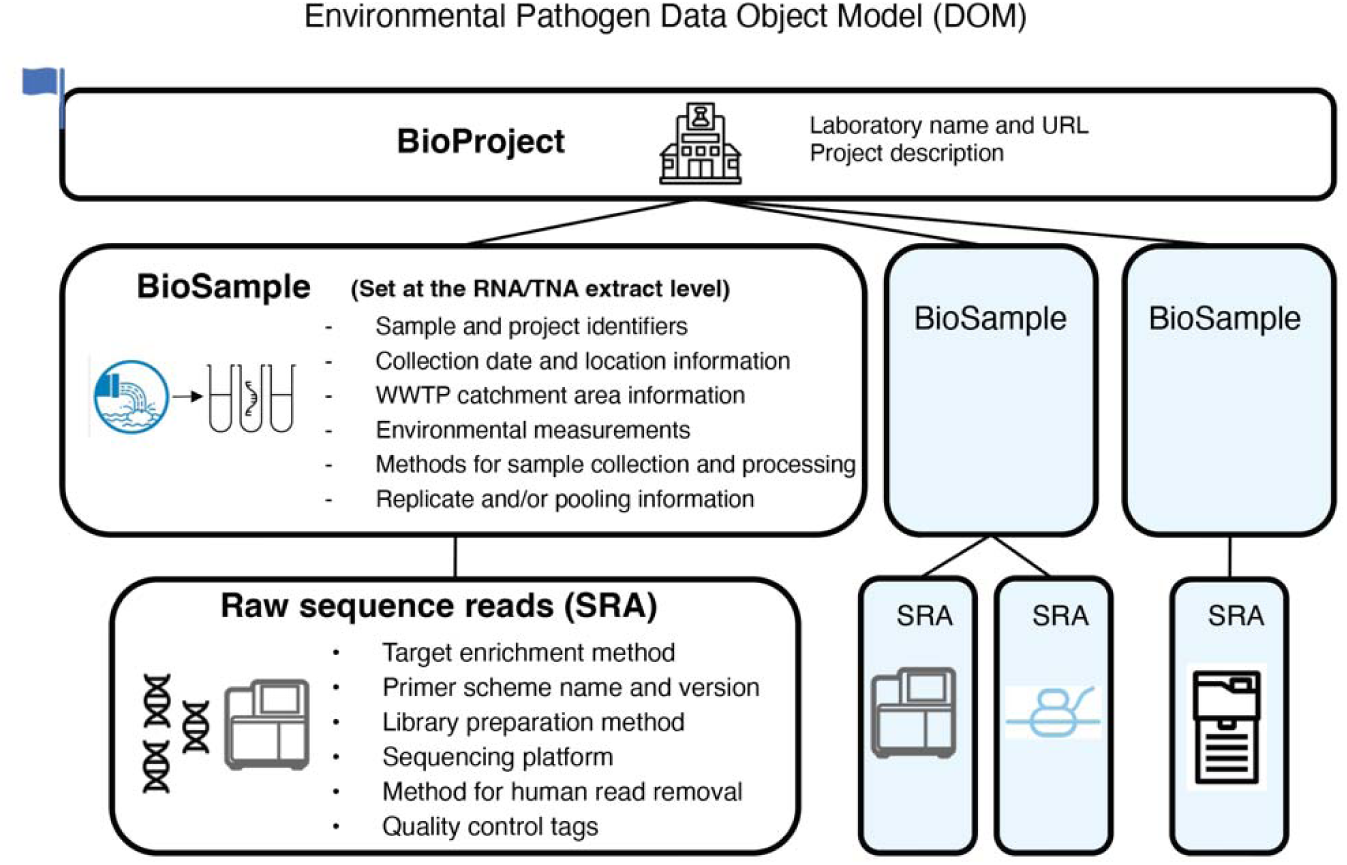
NCBI data structure for population-level pathogen surveillance, or Env. Pathogen Data Object Model (DOM). This Env pathogen DOM has sample and sequence contextual data required for analyzing wastewater sequence data with a single target pathogen, SARS-CoV-2. The flag on the BioProject represents the automated human-read scrubbing by NCBI for all data submissions linked to this project.

Our data package needed to include several key pieces of contextual data not previously included in NCBI’s BioSample SARS-CoV-2 wastewater template (30) or in their generic SRA metadata template. To fill this gap, we re-used fields from other packages where possible, including 1) sample-level pooling and replicate information, 2) sequence-level methods used for the targeted amplification of SARS-CoV-2, (31), and 3) known QC information as determined by the submitter (32). New custom attributes were created where needed to capture sample collection information (collection_time, collection_volume, instantaneous_flow, and collection_site_id) and laboratory methods for sequencing (enrichment_kit).

|  |  |
| --- | --- |
| collection_time* | For grab samples: the time of day the sample was collected in your timezone, 1-12 AM - 1-12 PM. |
| specimen_processing | Replicate and/or pooling information, critical for interpreting results |
| specimen_processing_id | Identifier used to track replicates and/or pooled samples |
| specimen_processing_details | Description of the experimental design, describing the technical or biological replicates and/or pooling design. |
| collection_site_id* | ID that uniquely identifies the sample collection site among other sample collection sites in this BioProject. It must be unique at the level of the submitter's data BioProject. Where possible, and with agreement from the facility, include the full name of the wastewater treatment plant. If anonymity is requested, create a masking ID to use for all samples collected at this site (e.g. AL-plant-1). |
| project_name | A concise name that describes the overall project or name of the coordinated sequencing effort from which the sequencing was organized. |
| collection_volume* | The volume of the sample collected, in mL |
| concentration_method* | The method used to concentrate a target organism, nucleic acid, or organelle within a sample. |
| extraction_method* | The protocol used to extract nucleic acids (DNA, RNA, or TNA) from a sample |
| extraction_control* | Organism (or nucleic acid) used in the extraction protocol to determine successful extraction. |
| instantaneous_flow* | For grab samples: enter the instantaneous volumetric flow measurement. The time of this measurement should correspond to when the grab sample was taken, and should be reported in UNITS of LITER per day. |
| **Additional SRA metadata attributes** |  |
| enrichment_kit* | Method used to enrich the target pathogen(s) |
| amplicon_PCR_primer_scheme | Provide the name and version of the primer scheme used to generate the amplicons for sequencing. |
| library_preparation_kit | Library preparation method used to convert a set of amplicons into a library ready for sequencing. |
| quality_control_method | Populate with the name of the method or pipeline used to evaluate sequence quality, often called "QC pipeline". |
| quality_control_method_version | List the version number of the quality control pipeline or method used. |
| quality_control_determination | Result of the quality control assessment. Leave blank if pass/fail thresholds have not been established or choose to flag an issue if known. |
| quality_control_issues | If there's a known or suspected quality control issue present in the sequence, choose from the available picklist to flag the issue. |
| quality_control_details | Free text space to include additional description of the flagged quality control issue. |
| dehosting_method | The method used to remove host reads from the pathogen sequence. |
| sequence_submitter_contact_email | Email contact for the lab that sequenced the isolate. |
| raw_sequence_data_processing_method | The method used for raw data processing such as removing barcodes, adapter trimming, filtering etc. |
| *custom attributes created for this project |  |

After the data structure was defined, we published an NCBI submission protocol adhering to this structure that included the custom BioSample and SRA metadata templates, capturing the full suite of contextual data needed for this project (33).

### Data flow and visualization

To effectively communicate and visualize the evolving landscape of SARS-CoV-2 variants detected in wastewater sites throughout the course of our project, we constructed an interactive dashboard in Tableau Desktop (Tableau Software LLC, Seattle, WA). Tableau offered a user-friendly interface, a broad set of dashboard design and development features, and could easily integrate multiple data sources, such as cloud queries of NCBI tables and output files from variant analysis pipelines.

The public dashboard needed to integrate several sources of data to present an informative snapshot of the project’s progress (Figure 2). NCBI Entrez queries summarized BioSample records without sequence data. Amazon Web Services (AWS) Athena queries of the SRA metadata table summarized metadata attached to raw sequence (SRA) and BioSample records. New sequence submissions under the BioProject PRJNA757291 were downloaded daily and analyzed with CFSAN’s Wastewater Analysis Pipeline (C-WAP) (34) for both QC metrics and to infer relative abundances of SARS-CoV-2 lineages in each sample, computed using the Freyja method (8). A static list of BioProjects, laboratory names, and wastewater sites served to organize the records recovered through NCBI and aid with the final dashboard visualizations.

**Figure 2.**
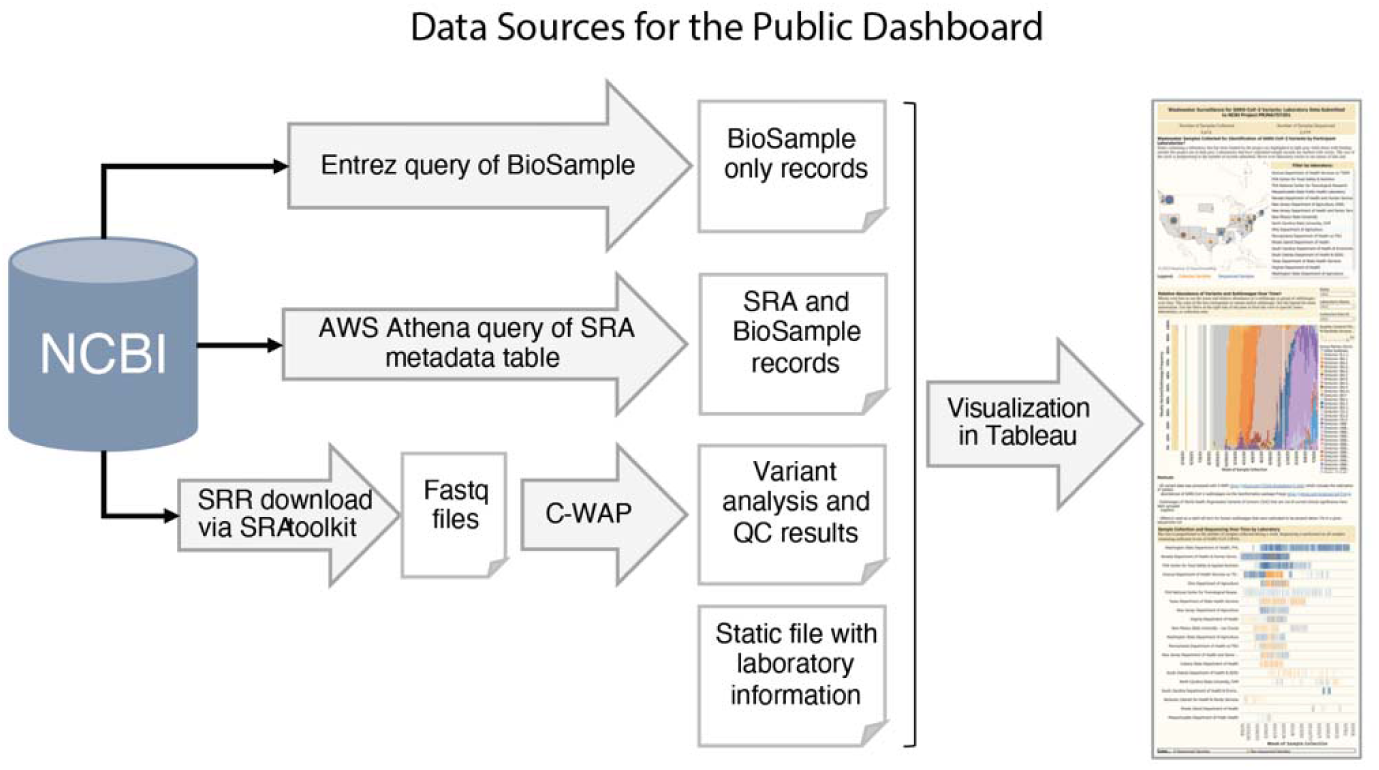
Data Sources for the public dashboard summarizing Wastewater Surveillance for SARS-CoV-2 Variants. The compilation of information for the public dashboard involved two distinct NCBI queries and a sequence analysis pipeline. Daily queries were executed to capture new submissions, and the newly obtained summary data were incorporated into the public dashboard guided by information in the static file. Raw data for dashboard available here: https://github.com/CFSAN-Biostatistics/WW-SC2-variant-estimations.

An important goal of the dashboard was to identify key aspects of the project that would be important to public health, such as geographic regions, stakeholders, temporality, sampling and sequencing progress, and variant calling. A map was used to display geography, stakeholders, and sampling and sequencing progress aspects to help communicate the scale of the project and the number of participating labs. A bar graph was used to show which SARS-CoV-2 variants were detected week to week, along with their relative abundances. Finally, a Gantt chart displayed the progress of participating labs in sampling and sequencing their samples over time.

Users were encouraged to explore the data visualizations by using filters to select which details they most wanted to see. Users could filter the dashboard by state, laboratory, and wastewater collection site. A quality control filter, which defaults to 80% of SARS-CoV-2 genome uncovered, was added to the dashboard on the public-facing webpage for users to filter data by % genome uncovered.

### Protocol pilot exercise

As this project entailed building methods to support expanded wastewater surveillance for state public health laboratories, it was important to establish consistency in analyses performed across participating laboratories. At the start of this project, FDA distributed a set of raw wastewater samples to each funded laboratory. These samples served two purposes: 1) they provided an early, standardized set of samples laboratories could use to test new methods, and 2) sequence data collected from each laboratory helped FDA identify which methods met the quality control requirements for this project. FDA collected four large volumes of wastewater (Table 1), comprising two samples taken about a month apart, each with two pseudo-replicates (grab samples taken back-to-back from the same location at the WWTP). Each large-volume sample was then aliquoted into 800ml samples. The October 2021 samples (WPP-sample_SA-1.01, WPP-sample_SA-2.01) were then spiked with 10^6^ copies of wildtype SARS-CoV-2 reference RNA (ATCC Heat Inactivated 2019 Novel Coronavirus strain nCoV/USA/WA-1/2020 Part #VR-1986HK). All bottles were frozen at -80C and then shipped to the laboratories on dry ice (four 800mL samples in each shipment). Each laboratory was asked to sequence the population of SARS-CoV-2 from each of these samples using methods of their choice then to submit their resulting raw sequence and contextual data to NCBI.

**Table 1.** Wastewater Protocol Pilot exercise samples.

| BioSample<br>"sample_name" | Collection<br>date | WWTP location | Treatment |
| --- | --- | --- | --- |
| WPP-sample_B.01 | 2021-09-20 | Mobile, AL | Raw wastewater |
| WPP-sample_C.01 | 2021-09-20 | Mobile, AL | Raw wastewater |
| WPP-sample_SA-1.01 | 2021-10-21 | Pascagoula, MS | Raw wastewater spiked with wt SARS-CoV-2, 10<sup>6</sup> copies/800mL |
| WPP-sample_SA-2.01 | 2021-10-21 | Pascagoula, MS | Raw wastewater spiked with wt SARS-CoV-2, 10<sup>6</sup> copies/800mL |

## RESULTS

### Participating laboratories

Twenty GenomeTrakr laboratories plus the FDA-CFSAN laboratories received funding for this special project (Table 2). Each laboratory identified at least two wastewater sites (Supplemental Table 1) for routine sampling (1-2x times a week) for a minimum of six months. In total, samples from 81 sites were included in this project. Where feasible, sites were in counties with a higher relative percentage of food and agriculture workers. Sites included both municipal wastewater treatment plants and direct wastewater lines from food processing facilities – spanning both urban and rural populations.

**Table 2.** List of participating laboratories. Twenty GenomeTrakr laboratories plus FDA-CFSAN participated in this project.

| Laboratory names |
| --- |
| Arizona State Department of Health Services, T-Gen North |
| California Department of Public Health |
| Indiana State Department of Health |
| Kentucky State Cabinet for Health and Family Services |
| Massachusetts State Department of Public Health |
| Nevada State Public Health Laboratory, University of Nevada - Reno |
| New Jersey State Department of Agriculture |
| New Jersey Department of Health |
| New Mexico State University - Las Cruces |
| North Carolina State University - Raleigh |
| Ohio State Department of Agriculture |
| Pennsylvania State University - University Park |
| Rhode Island Department of Health, State Health Laboratory |
| South Carolina Department of Health and Environmental Control |
| South Dakota State University |
| Texas Department of State Health Services |
| Virginia Division of Consolidated Laboratory Services |
| Washington State Department of Agriculture |
| Washington State Department of Health |
| West Virginia Department of Agriculture |
| FDA-Center for Food Safety and Applied Nutrition |

### Wastewater Protocol Pilot exercise

Ten laboratories participated in a pilot exercise to access different laboratory methods being utilized: nine labs provided sequence data from all four distributed samples (Table 1). Those nine laboratories successfully amplified and sequenced SARS-CoV-2 from the four WPP samples (Supplementary Table 2) and submitted their resulting sequences to NCBI (BioProject, PRJNA767800). These submissions were obtained by a diverse array of methods: seven extraction methods, six concentration methods, four enrichment strategies, seven primer schemes, and seven library preparation methods. One remaining lab opted out of the sequencing portion of the exercise, as they had encountered issues with their ddPCR (droplet digital PCR) method.

The cumulative submissions from multiple laboratories totaled 17 datasets for the four samples. Although the limited number of replicates precludes drawing definitive conclusions about individual or combined methods, several overarching trends emerged that informed our subsequent decisions for real-time sampling. In particular, the “Percent reads aligned” metric confirmed the robust specificity of three different enrichment methods for the SARS-CoV-2 virus: QIAseq DIRECT SARS-CoV-2, NEBNext ARTIC SARS-CoV-2, and Illumina COVIDSeq. The “Percent SARS-CoV-2 genome covered” demonstrated the strong performance of most primer schemes accessed in this exercise. Furthermore, across all tested methods, the variant analyses were largely consistent across samples. Specifically, the sequences for “WPP-sample_B.01” and “WPP-sample_C01” showed mostly Delta variants, while “WPP-sample_SA-1.01” and “WPP-sample_SA-2.01” revealed strong wild-type signal originating from the spiked-in synthetic virus (Supplementary Figure 1).

### Quality control thresholds

We determined approximate QC thresholds for the sequence data based on data collected from the protocol pilot exercise and from the first couple months of sequencing. Important considerations for setting these thresholds included determining the percentage of the SARS-CoV-2 genome coverage required to confidently identify population-level variants and sub-lineages, the necessary depth of coverage to capture most of the circulating lineages, the percentage of SARS-CoV-2 in the raw sequence data, and the identification and removal of human sequencing reads prior to public release. We described four QC bins that capture major categories of sequence quality (Table 3) and proposed QC thresholds for three metrics we identified as important for determining high-quality data: % SARS-CoV-2 reads, % SARS-CoV-2 genome uncovered, and average genome coverage depth.

**Table 3.** Quality control (QC) for SARS-CoV-2 sequencing data. Four QC categories, or bins, were established based on thresholds set for various QC metrics. Sequence tags were developed to communicate these categories directly on the sequence file in an attribute called “quality_control_determination”. Only data with QC tagged in the A or B bin was visualized on the public FDA variant analysis dashboard.

| QC bin | QC bin description | % genome uncovered (<10X) | Average coverage | Other observations | Submit to NCBI | Tag for SRA attribute: "quality_control_determination" | Included in FDA Dashboard |
| --- | --- | --- | --- | --- | --- | --- | --- |
| A | No QC issues evident | <5% | >1000X | >50% reads are SARS-CoV-2 | Yes | no quality control issues identified | Yes |
| B | Some QC issues, but variant calling likely OK | 6% - 40% | 100X-1000X |  | Yes | minor quality control issues identified | Yes |
| C | Insufficient data for confidence in variant calling | 40% - 95% | 10X-100X | low fraction of lineage specific mutations, (C-WAP reports) | Yes | sequence flagged for potential quality control issues | No |
| F | Significant QC and/or study design issues | >95% | < 10X | < 5% reads SARS-Cov-2, suspected contamination (SNR low), low sequence quality, etc. | No | sequence flagged for significant quality control issues | No |

Once we had a QC target for sequence data, we identified three critical QC checkpoints in laboratory workflow (Figure 3). *QC check #1*: Samples containing no detectible SARS-CoV-2 RNA were deemed to have failed QC and were not processed further. However, samples that contained any level of target RNA, even at very low levels, were considered “passing” and sent on to the cDNA synthesis step for amplification. *QC check #2*: To determine whether investing time in library preparation and sequencing was justifiable in terms of cost and effort, a thorough quality assessment of the PCR product (targeted enrichment) was conducted using the Qubit HS kit and a fragment size analyzer, such as Agilent Tape Station or Bioanalyzer (24, 26). Samples passing this QC step were selected for sequencing. *QC check #3*: The final major QC check involved a rigorous evaluation of the raw sequencing data. For this purpose we developed SSQuAWK4 (35), to automate the QC process for our laboratories; this tool was then made publicly accessible through a custom Galaxy instance, GalaxyTrakr (36). We also used a thorough QC evaluation and variant calling pipeline, CFSAN Wastewater Analysis Pipeline (C-WAP), via command line interface (34). Both reports included key summary metrics, such as percentage of total reads aligned to the SARS-CoV-2 reference genome, average depth of coverage, and percentage of the SARS-CoV-2 genome uncovered (<10X).

**Figure 3.**
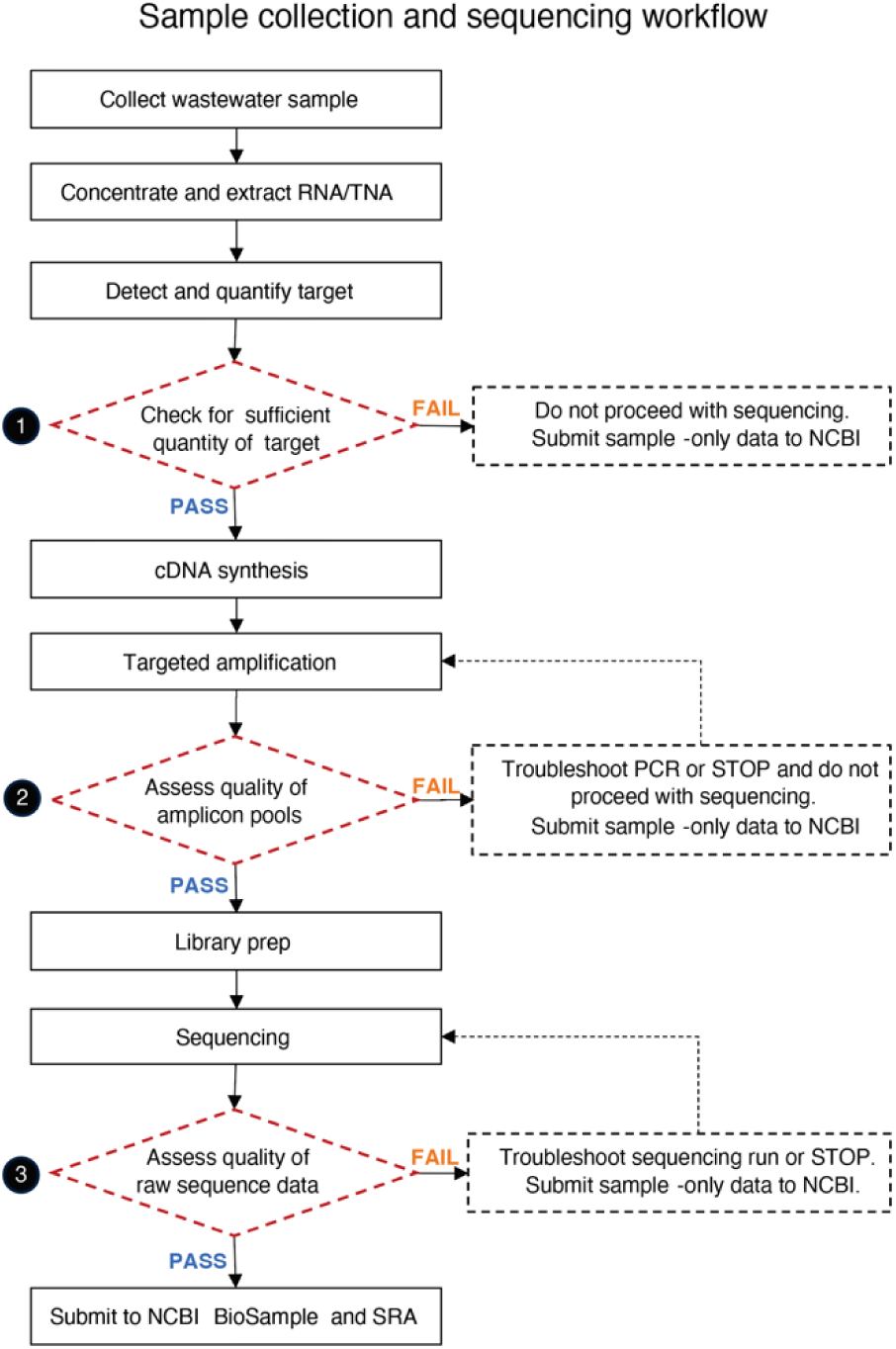
Sample collection and sequencing workflow. SARS-CoV-2 wastewater surveillance sample analysis process and recommended quality control checkpoints used in this project.

Based on that QC assessment, each sequence was assigned a QC bin (A, B, C, or F). While we performed QC assessments from very beginning of data collection, there was some uncertainty about what would qualify as a “high” or “low” quality sequencing run. Therefore, we submitted almost all sequence data to NCBI’s Sequence Read Archive (SRA), along with metadata and QC evaluations (32). Then, to guide future laboratory practices, we devised a decision matrix (Table 3), which became instrumental for downstream selections of which samples merited being featured on the public-facing dashboards.

### Summary of data collected

Routine, systematic random, wastewater sample collection for this project was initiated in September 2021 and ended by June 2023, with contributing laboratories submitting sequences in staggared 6-month time periods (Supplemental Figure 2). When detectable levels of SARS-CoV-2 were present determined by quantification of COVID-specific RT/dd -PCR targets as a first phase screening, targeted amplicon approaches were used to sequence the SARS-CoV-2 RNA in the sample. In total, 3,406 wastewater samples were collected, of which 2,517 were subjected to sequencing. The resulting raw sequence data and comprehensive set of standard contextual data were submitted to laboratory-specific NCBI BioProjects, nested under the umbrella BioProject PRJNA757291. Every sample collected and tested for this project has a BioSample entry, even the ones that were not sequenced, thereby providing a unique dataset within NCBI that includes both positive and negative samples.

Standard terminology describing sample collection and sequencing methods were included as attributes on both the sample record (BioSample) and and experiment records (SRA submission). Sample processing methods utilized varied across the project (Table 4). There was good representation of composite versus grab samples, n=2023 (59.7%) and n=1466 (40.1483 (3%), respectively. Most labs collected raw wastewater, n=3305 (97.5%), with a few primary effluent and post-grit removal samples also included. Among the concentration methods used, 90% of samples were concentrated using one of five methods: Ceres Nanotrap (n=1212), Innovaprep ultrafiltration (n=604), Promega large volume TNA capture kit (n=455453), PEG (polyethylene glycol) precipitation + ultracentrifugation (n=420), and Centricon 100k (n=357). For nucleic acid extraction, the most employed method was the Qiagen MagMAX Viral Kit (n=939, 28%), followed by a variety of similiar Promega extraction kits (n=627, 19%).

**Table 4.** Sample methods included on NCBI’s public BioSample records. Methods cover type of sample collection, wastewater sample matrix, viral concentration method, and nucleotide extraction method.

| Sample Type | # of BioSamples | # of Labs | % of Total |
| --- | --- | --- | --- |
| composite | 2023 | 18 | 59.7 |
| grab | 1466 | 8 | 40.3 |
| Total | 3389 |  |  |
| **Sample Matrix** | **# of BioSamples** | **# of Labs** | **% of Total** |
| Raw Wastewater | 3306 | 19 | 97.5 |
| Post-Grit Removal | 79 | 1 | 2.3 |
| Primary Effluent | 4 | 1 | 0.1 |
| Missing | 1 | 1 | <0.1 |
| Total | 3389 |  |  |
| **Viral Concentration Method** | **# of BioSamples** | **# of Labs** | **% of Total** |
| Ceres Nanotrap | 1212 | 8 | 35.8 |
| Innovaprep ultrafiltration | 604 | 5 | 17.8 |
| Promega wastewater large volume TNA capture kit | 455 | 5 | 13.4 |
| Peg precipitation + ultracentrifugation | 420 | 4 | 12.4 |
| Centricon 100k | 357 | 1 | 10.5 |
| Zymo Water Concentration Buffer | 155 | 1 | 4.6 |
| Skim Milk Flocculation | 99 | 1 | 2.9 |
| Membrane filtration with acidification and MgCl2 | 64 | 2 | 1.9 |
| Innovaprep CP Select | 19 | 1 | 0.5 |
| backflushed raw ww using rexseed filters; Promega wastewater large column TNA capture kit | 4 | 1 | 0.1 |
| Total | 3389 |  |  |

| **Nucleotide Extraction Method** | **# of BioSamples** | **# of Labs** | **% of Total** |
| --- | --- | --- | --- |
| Qiagen MagMAX Viral Kit | 939 | 2 | 27.7 |
| Promega Extraction Kits | 627 | 5 | 18.5 |
| Qiagen AllPrep Powerviral DNA/RNA kit | 424 | 4 | 12.5 |
| Qiagen RNeasy Powerwater Kit | 324 | 4 | 9.6 |
| Zymo quick-rna viral kit | 345 | 2 | 10.2 |
| Zymo Environ Water RNA Kit (R2042) | 262 | 1 | 7.7 |
| QIAamp Viral RNA mini kit | 201 | 5 | 5.9 |
| neb monarch total rna miniprep kit + zymo onestep pcr inhibitor removal kit | 155 | 1 | 4.6 |
| mcaherey-nagel nucleomag DNA/RNA water kit | 108 | 1 | 3.2 |
| Ceres Nanotrap | 4 | 2 | 0.1 |
| Total | 3389 |  |  |

Sequencing methods were captured at the experiment-level, attached to the raw sequence data (Table 5). Target enrichment methods encompassed broad categories of tiled amplicon approaches, with 90% of submissions choosing QIAseq DIRECT (n=923, 37%), NEBNext ARTIC (n=825, 33%), or the Illumina COVIDSeq Assay (n=433, 17%). PCR primer schemes for these enrichment approaches evolved alongside the virus – in total there were 11 different primer schemes used across the project. Seven different library preparation kits were utilized to prepare the SARS-CoV-2 amplicons for sequencing and 10 different sequencing platforms were used to generate sequence data. Illumina instruments comprised 90% of the sequences submitted (n=2257), followed by Oxford Nanopore Technology (ONT) (n=256, 10%), and finally, a small number sequenced on a PacBio instrument (n=4).

**Table 5.** Sequencing methods included on NCBI’s public SRA records. Methods cover enrichment kit (general approach for enriching the target pathogen), Amplicon PCR primer scheme, library preparation kit, and sequencing instrument name.

| **Enrichment Kit** | **# of Sequences** | **# of Labs** | **% of Total** |
| --- | --- | --- | --- |
| QIAseq DIRECT SARS-CoV-2 | 923 | 3 | 36.7 |
| NEBNext ARTIC SARS-CoV-2 RT-PCR Module | 825 | 12 | 32.8 |
| Illumina COVIDSeq Assay | 433 | 1 | 17.2 |
| Swift Normalase Amplicon SARS-CoV-2 Panels | 199 | 1 | 7.9 |
| Not Applicable | 137 | 4 | 5.4 |
| Total | 2517 |  |  |
| **Amplicon PCR Primer Scheme** | **# of Sequences** | **# of Labs** | **% of Total** |
| ARTIC V3 | 125 | 1 | 5.0 |
| ARTIC V4 | 308 | 1 | 12.2 |
| ARTIC V4.1 | 131 | 4 | 5.2 |
| NEB VarSkip 1a Long | 18 | 1 | 0.7 |
| NEB VarSkip 1a Short | 125 | 5 | 5.0 |
| NEB VarSkip 2a Short | 307 | 8 | 12.2 |
| NEB VarSkip 2b Short | 369 | 7 | 14.7 |
| QIAseq DIRECT SARS-CoV-2 - Boosted | 97 | 2 | 3.9 |
| QIAseq DIRECT SARS-CoV-2 Primers | 838 | 3 | 33.3 |
| SARS-CoV-2 SNAP primer pool | 199 | 1 | 7.9 |
| Total | 2517 |  |  |
| **Library Preparation Kit** | **# of Sequences** | **# of Labs** | **% of Total** |
| Amplicon Sequencing Kit (PacBio) | 4 | 1 | 0.2 |
| Illumina DNA Prep | 513 | 8 | 20.4 |
| Ligation sequencing kit | 286 | 3 | 11.4 |
| NEBNext ARTIC SARS-CoV-2 Library Prep Kit (Illumina) | 514 | 5 | 20.4 |
| NEBNext Ultra II FS DNA Library Prep for Illumina | 6 | 1 | 0.2 |
| QIAseq DIRECT Unique Dual Index Prep | 995 | 4 | 39.5 |
| Swift Normalase Amplicon SARS-CoV-2 Panels | 199 | 1 | 7.5 |
| Total | 2514 |  |  |
| **Sequencing Instrument** | **# of Sequences** | **# of Labs** | **% of Total** |
| Illumina iSeq 100 | 55 | 2 | 2.2 |
| Illumina MiniSeq | 440 | 4 | 17.5 |
| Illumina MiSeq | 1622 | 14 | 64.4 |
| Illumina NovaSeq 6000 | 83 | 1 | 3.3 |
| NextSeq 550 | 57 | 1 | 2.3 |
| GridION | 8 | 1 | 0.3 |
| MinION | 248 | 1 | 9.9 |
| Sequel II | 4 | 1 | 0.2 |
| Total | 2517 |  |  |

### Quality of sequence data

As could be expected from a multi-laboratory project using various field sampling approaches, and performing simultaneous methods development and data collection, the quality of sequences submitted for this project exhibited significant variability, ranging from exceptional to very low quality, based on the predefined thresholds outlined in Table 3. Of the 2,255 Illumina short read sequences, 1381 (61%) were categorized as having "no quality control issues" (A bin), 219 (10%) as "minor quality control issues" (B bin), 633 (28%) as having "potential quality control issues" (C bin), and 22 (<1%) were flagged for "significant quality control issues" (F bin). For the average depth of SARS-CoV-2 genome coverage thresholds, we targeted 1,000X as an ideal, while considering 100X as the minimum. Similarly, the submitted sequence data ranged widely from less than 10X to over 135,000X (Figure 4a), with a notable concentration of sequences below these thresholds flagged as low quality (Figure 4a). Percent of genome uncovered (i.e., percent of the SARS-CoV-2 genome not sequenced with at least 10X coverage) showed a similar pattern, with 72% of submissions falling under our threshold of 40% (Figure 4b).

**Figure 4.**
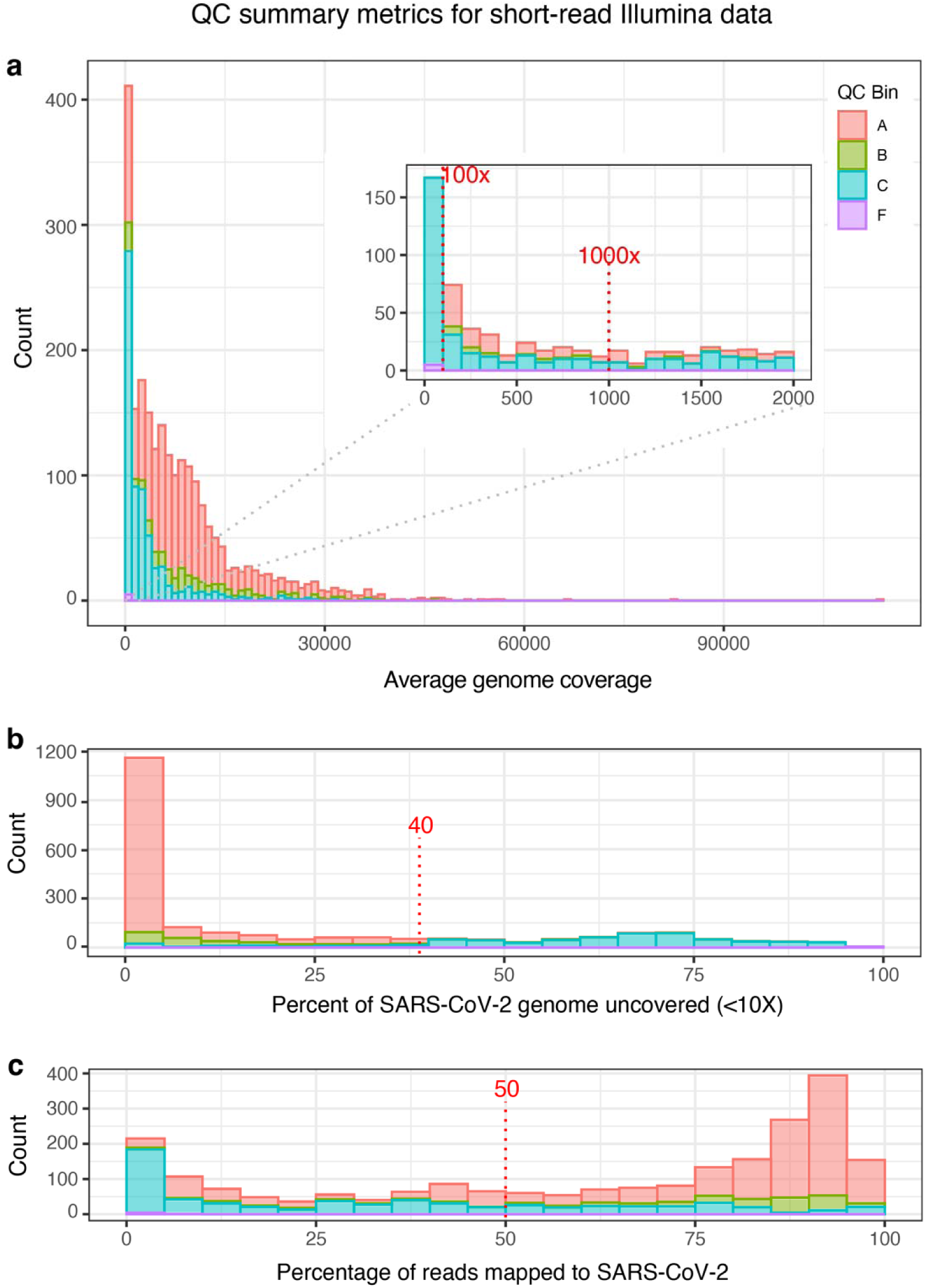
QC summary metrics for short-read Illumina data. Three panels summarize the quality of population-level SARS-CoV-2 sequence data collected and submitted for this project: **a)** Average depth of coverage across the SARS-CoV-2 genome (average coverage), **b)** Percent of the SARS-CoV-2 genome uncovered at <10X, and **c)** Percent of raw sequence reads that aligned to the SARS-CoV-2 genome. Quality control determinations made by the submitter (QC bins A, B, C, or F) are also summarized in each panel.

Sequences for which more than 40% of the genome had not been sequenced were predominantly flagged into the C and F bins (Figure 4b). Conversely, submissions under this threshold, for which more of the genome had been successfully sequenced, were mostly assigned an A or B bin, indicating the coverage of the SARS-CoV-2 genome was suitable for variant analysis. Finally, for each submission, we computed the percentage of raw sequence reads that mapped to the SARS-CoV-2 genome. We see a general trend of reads with high percentages of SARS-CoV-2 being higher quality (Figure 4c) and, conversely, reads with lower percentages of SARS-CoV-2 having a lower QC assessment, although there is no obvious inflection point at 50%, which was our target goal. We had plenty of sequences categorized as high quality, in Bin A, even though only a small fraction of reads might have been identified as SARS-CoV-2.

### Variant analysis

To visualize the variants and sub-lineages in wastewater samples over time, we plotted their relative abundance against week of collection, from September 12, 2021 through June 4, 2023 (Figure 5)(37). The dashboard was updated when new submissions appeared at NCBI, with a maximum frequency of once per day, aiming for current data representation. Analysts also continuously monitored public health news for mentions of new and clinically important variants that should be added to the dashboard’s legend. Sub-lineages that were not of public health importance or did not contribute more than 1% relative abundance within each sample were collectively categorized as “Others” for the purposes of the public dashboard.

**Figure 5.**
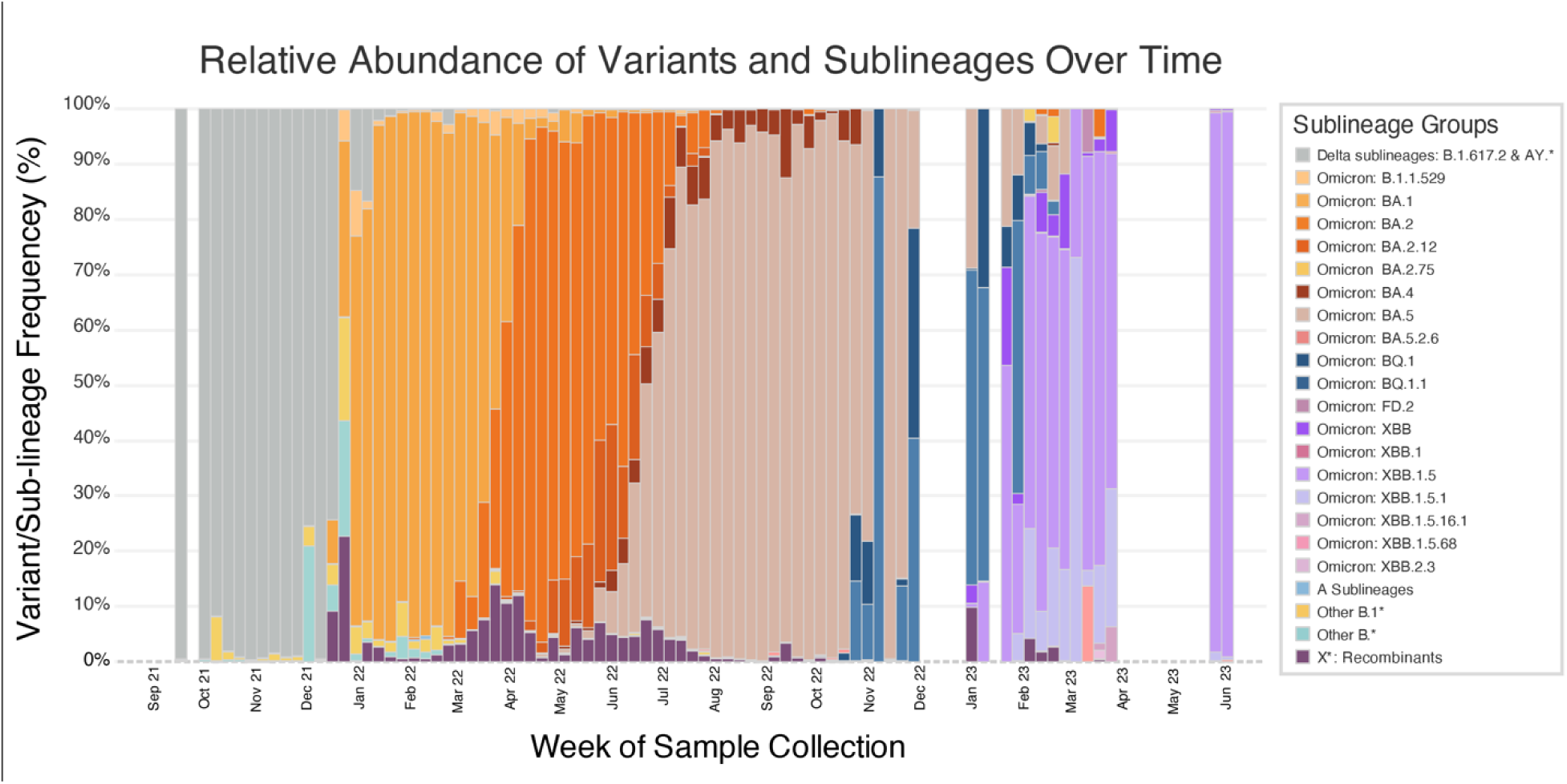
Relative abundances of variants and sublineages over time. Stacked bar chart showing the average variant and sub-lineage proportions for samples collected during that week. For the sake of visibility, only sub-lineages with a relative abundance of ≥5% for at least one week are displayed. The rest, regardless of its designated interest to the WHO or CDC, were treated as parts of their parent lineage until a sub-lineage had sufficient relative abundance to meet the ≥5% threshold.

Across the contributing laboratories, most of the sampling occurred in in 2022 (Supplemental Figure 2), resulting in a few dashboard gaps in late 2022 and 2023 (Figure 5). Samples collected from September 2021 through early December 2021 all belonged to Delta sub-lineages. Omicron BA.1 made its initial appearance during the week of December 12th, 2021, swiftly replacing nearly all circulating Delta lineages within the subsequent month. Following this, Omicron BA.2 was identified in our samples in mid-March 2022, taking over from BA.1 by the end of April 2022. In early May, Omicron BA.4 emerged and circulated until October 2022, although it never reached dominance. In late April 2022, Omicron BA.5 was detected and became the predominant circulating sub-lineage until October 2022, when Omicron BQ lineages started appearing. The first widely circulating hybrid Omicron lineage, XBB, emerged in November 2022 and maintained dominance through June 2023.

### Turnaround Time

In line with our project’s primary objective of delivering timely pandemic sequence data for public health purposes, we evaluated the turnaround time (TAT) as the number of days from sample collection to NCBI data release for each participating laboratory. Our analysis revealed two distinct categories of laboratories based on their approach to sample processing (Figure 6). The first category consisted of five laboratories, including FDA, that processed samples as they were collected, resulting in an average TAT range of approximately 15-30 days. The second category comprised 12 laboratories that initially collected samples but processed them at a later date due to various factors, including supply-chain delays for reagents and instruments, hesitancy within state public health laboratories to publically release data that were collected using non-validated methods (e.g. Lab P), and staffing shortages due to pandemic response burden. Within this category, the average TAT exhibited significant variation, ranging from 60 to 410 days between sample collection and data submission.

**Figure 6.**
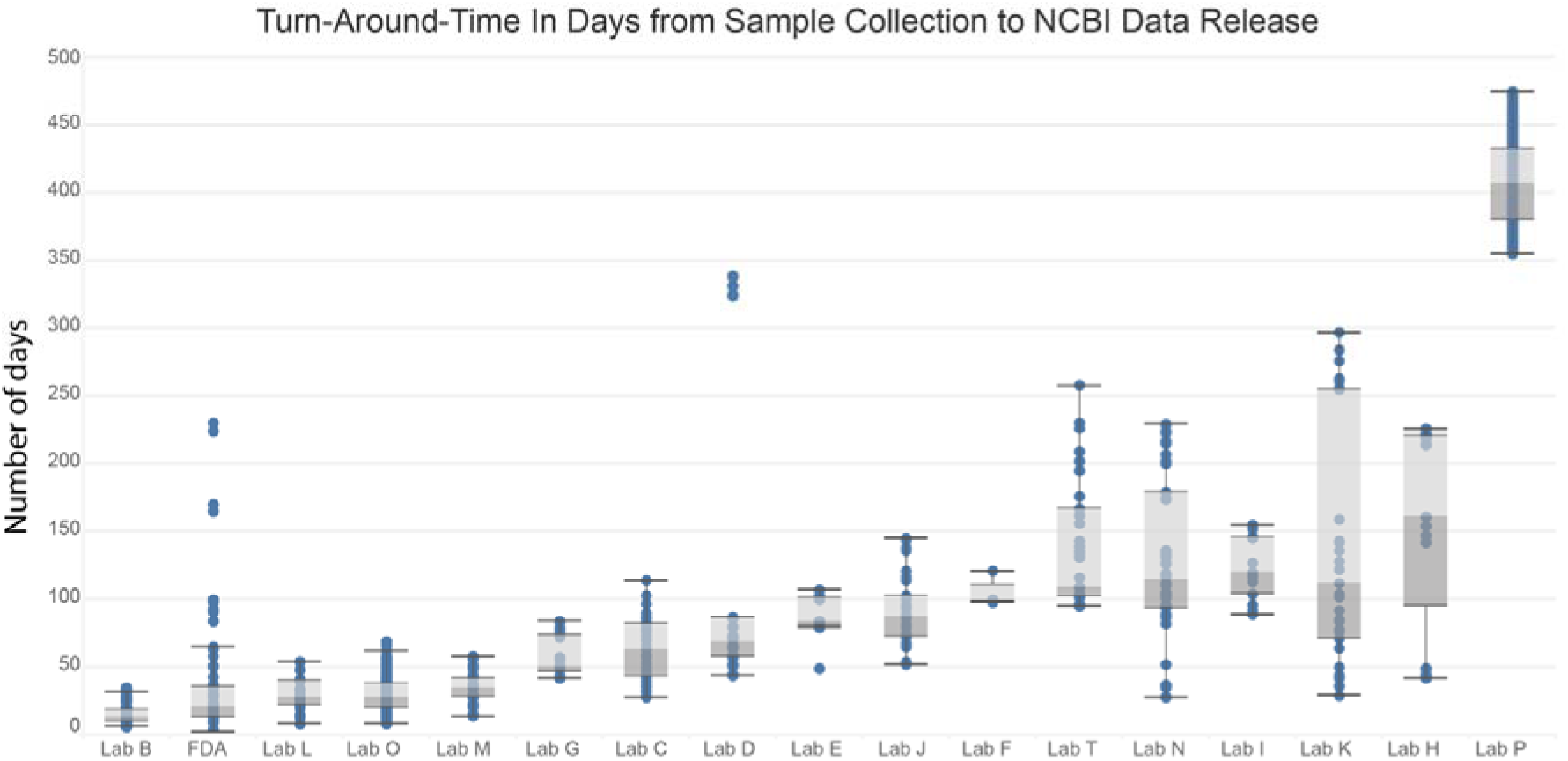
Turnaround Time from Sample Collection to NCBI Data Release. Box and whisker plot showing number of days between sample collection date and NCBI release date for each participating laboratory.

## DISCUSSION

This project represents the first nation-wide, culture-free, population-level surveillance of a pathogen, with the intention to make data publicly available as it was collected. Within six months of funding acquisition, the GenomeTrakr program successfully implemented surveillance for a novel pathogen, sourced from a new-to-the-program sample origin, despite the requirement for developing new sample collection and preparation methods, optimization of novel sequencing and analysis methods, and need for novel contextual data fields. We demonstrated that these methods work and multiple U.S. local public health, agriculture, and academic laboratories within our network are now equipped and trained to execute these methods when requested at short notice. Data generated through these accomplishments underscore the enduring potential of wastewater sampling as an emerging surveillance tool.

Though largely successful, we encountered several challenges inherent to the targeted amplicon approach chosen for sequencing SARS-CoV-2 in the samples. Our initial primer sets for the targeted amplicons had been designed on previously circulating lineages of SARS-CoV-2; however the ongoing evolution of the SARS-CoV-2 genome during multiple Omicron waves (BA.2, BA.4, BA.5) (38) resulted in periodic dropouts in coverage, or primer pairs that would suddenly stop working. Minor updates to the primer schemes were released in response (39, 40), however these needed to be verified internally to to ensure they worked before we recommend their adoption across our network of laboratories. This proved demanding to keep pace with, necessitating continuous evolution of protocols and metadata template updates alongside our routine surveillance efforts.

Due to their intrinsic reliance on the external sources regarding all SARS-CoV-2 variants ever reported in the literature, similar adaptations were required to ensure bioinformatic analyses were robust and the data analysis pipelines remained current. Each time a new variant or sub-lineage of significance was named, the variant database needed to be updated and the entire dataset feeding the dashboard required re-analysis. This dynamic stands in stark contrast to the WGS protocol employed over the past decade (41), where a consistent protocol works reliably for all enteric bacterial pathogens, and updates to that protocol are infrequent occurrences.

As a direct result of constantly updating laboratory methods, there were periods of time when we were not confident in the variant calling until we were sure participant labs had implemented the primer updates. For example,, as the virus mutated further in early 2022, multiple “Omicron” lineages were co-circulating, resulting in some lineages only differing by a few loci, further compounded by multiple other mutations evolving under convergent evolution (42). If the sequencing missed one or more of these diagnostic loci due to a now-suboptimal experimental design, we would expect an over-representation of parent lineages, mirrored by under-representation of the true variant(s). To address this problem, we attempted to use the QC flags to communicate how confident we were with our sequencing data.

Despite those challenges, wastewater is an ideal environmental sample to target for this project because it captures pathogen shedding at the population or subpopulation level within a spatially explicit geographic region (sewershed or subsewershed). Unlike well-established WGS-based surveillance systems (3, 4), the SARS-CoV-2 amplicon-based sequencing approach requires no culturing step, shaving days to weeks off the turnaround time from sample collection to acquiring sequencing results. For this reason, this project met two important goals for FDA’s GenomeTrakr program: 1) to contribute timely genomic data for SARS-CoV-2 pandemic response, and 2) to develop capacity and best practices for culture-independent, population-level, environmental surveillance for other pathogens of interest to the FDA, namely enteric pathogens central to our food safety mission.

Drawing from our success in managing a laboratory network funded to sequence pure-culture enteric pathogens isolated from environmental and other non-human sources, with NCBI serving as our primary repository (4), we propose the following best practices for employing a comparable distributed laboratory model and utilizing NCBI as the primary repository for the implementation of culture-independent, population-level sequencing of a pathogen from wastewater. 1) Establish a standard data structure, or Data Object Model (DOM), within NCBI (or other repository within the International Nucleotide Sequence Database Collaboration, INSDC) to capture the sequence data and large suite of contextual data. 2) Create a custom FAIR contextual data standard that captures relevant sample and sequence metadata, maps to the DOM, and is interoperable with existing INSDC standards. 3) Define the critical steps within the methods for assessing QC and set thresholds for determining next steps. 4) Publish version-controlled protocols that cover delineation of sewersheds/subsewersheds, sample collection, laboratory methods, quality control assessment, analysis, and INSDC data submission. 5) Process, sequence, and upload sample data to support timely public health actions (not entirely met by our project, but recommended for future efforts). Lastly, 6) develop a public dashboard to visualize current data collection and analysis results to serve the needs of the project.

## Data Availability

SARS-CoV-2 sequence data + contextual data collected through this project are available under the NCBI BioProject, PRJNA757291. Variant analysis results are available through links posted on our dashboard: https://www.fda.gov/food/whole-genome-sequencing-wgs-program/wastewater-surveillance-sars-cov-2-variants or through.

https://www.ncbi.nlm.nih.gov/bioproject/PRJNA757291

## Acknowledgements

This project was supported in part by funding from the American Rescue Plan Act of 2021 and an appointment to the Research Participation Program at the U.S. Food and Drug Administration administered by the Oak Ridge Institute for Science and Education through an interagency agreement between the U.S. Department of Energy and the U.S. Food and Drug Administration. We would like to acknowledge John Callahan and CFSAN senior leadership for support on this project; Lili Velez for scientific editing; Sebastian Cianci and the web team for help getting the dashboards published; Justin Payne for advice on maintenance and scalability of software; Amy Kirby and Rory Welsh at CDC’s National Wastewater Surveillance System for collaboration; Arvind Varsan at Arizona State University for early discussions on sequencing methods; Rose Kantor and Stacia Wyman at UC Berkeley for early discussions on sequencing methods; Jay Garland at EPA and Seth A. Faith at Ohio State University for early discussions on sequencing strategy; Volodymyr Tryndyak and Camila Silva from FDA’s National Center for Toxicological Research for collaborative discussions; FDA’s 21Forward team for help with maps to help choose the wastewater sites; Josh Levy at the Scripps Research Institute: La Jolla, CA, for advice using Freyja; and Rick Lapoint, John Anderson and the NCBI SRA and BioSample teams for handling all our curation requests.

## Supplemental material

Supplemental Figure 1 – WPP variant analysis

Supplemental Figure 2 – Gantt chart

Supplemental Table 1 – Wastewater sites

Supplemental Table 2 – WPP data summary

Supplemental File 1 – BioSample ww template

Supplemental File 2 – SRA metadata template

## GenomeTrakr consortium authors

Ward Jacox, Dave Engelthaler, Michael Valentine, Crystal Hepp from Arizona State Department of Health Services and TGen; David Kiang and Zhirong Li from California Department of Public Health; Ryan Gentry, Mary Ann Hagerman, Mary Robinson, Jesse Knibbs, Madi Asbell from Indiana State Department of Health; Beth Johnson, Logan Burns, Ashley Aurand-Cravens, and Joshua Stacy from Kentucky State Cabinet for Health and Family Services; Tracy Stiles, Esther Fortes, Matthew Doucette, Brandon Sabina, Luc Gagne, and Kelly Binns from Massachusetts State Department of Public Health; Mark Pandori, Andrew Gorzalski, and Lauryn Massic from Nevada State Public Health Laboratory; Sarmila Dasgupta, Amar Patil, and Apryle Panyi from New Jersey Department of Agriculture, Animal Health Diagnostic Laboratory; Edward Acheampong, Thomas Kirn, and Nicholas Palmateer from New Jersey State Department of Health; Willis Fedio and Yatziri Preciado from New Mexico State University - Las Cruces, and Srikanth Paladugu from New Mexico Department of Health; Siddhartha Thakur, Lyndy Harden-Plumley, and Luke Raymond from North Carolina State University – Raleigh; Melanie Prarat, Ashley Sawyer, and Jonah Perkins from Ohio Department of Agriculture; Edward Dudley, Jasna Kovac, Nkuchia M. M’ikanatha, Erin M. Nawrocki, Yezhi Fu, and Nyduta Mbogo from Pennsylvania State University - University Park; Kristin Carpenter-Azevedo, Richard C. Huard, and Sean Sierra-Patev from Rhode Island Department of Health, State Health Laboratory; Megan Davis, Laura M. Lane, Christy A. Jeffcoat, Gregory Goodwin, Gabrielle Godfrey, Smith, Andrew, Chukwuemika N Aroh, Kirsti R. Gilmore, and Jessica Freeman from South Carolina Department of Health and Environmental Control; Joy Scaria, Jane Hennings, and Eric Nelson from South Dakota State University; Yan Sun, Bonnie Oh, and Michael Jost from Texas Department of State Health Services, and Bryan Brooks and Laura Langan from Baylor University, Waco, Texas; Lauren Turner, Stephanie Dela Cruz, Jessica Maitland, Shelby Bennett, Logan Fink, Mary Toothman, and Hyunsook Moon from Virginia Division of Consolidated Laboratory Services; Yong Liu and Mychal Hendrickson from Washington State Department of Agriculture; Darren Lucas, Phillip Dykema, Roxanne Meek, Geoff Melly, Paige Sickles, Breanna McArdle, and Anneke Jansen from Washington State Department of Health; Megan Young, Josh Arbaugh, and Zachary Kuhl from West Virginia Department of Agriculture; and Ewa King from the Association of Public Health Laboratories.

## Notes

### Competing Interest Statement

The authors have declared no competing interest.

