## Supplementary figures and images for "SARS-CoV-2 wastewater variant surveillance: pandemic response leveraging FDA’s GenomeTrakr network"

### Supplemental Figure 1

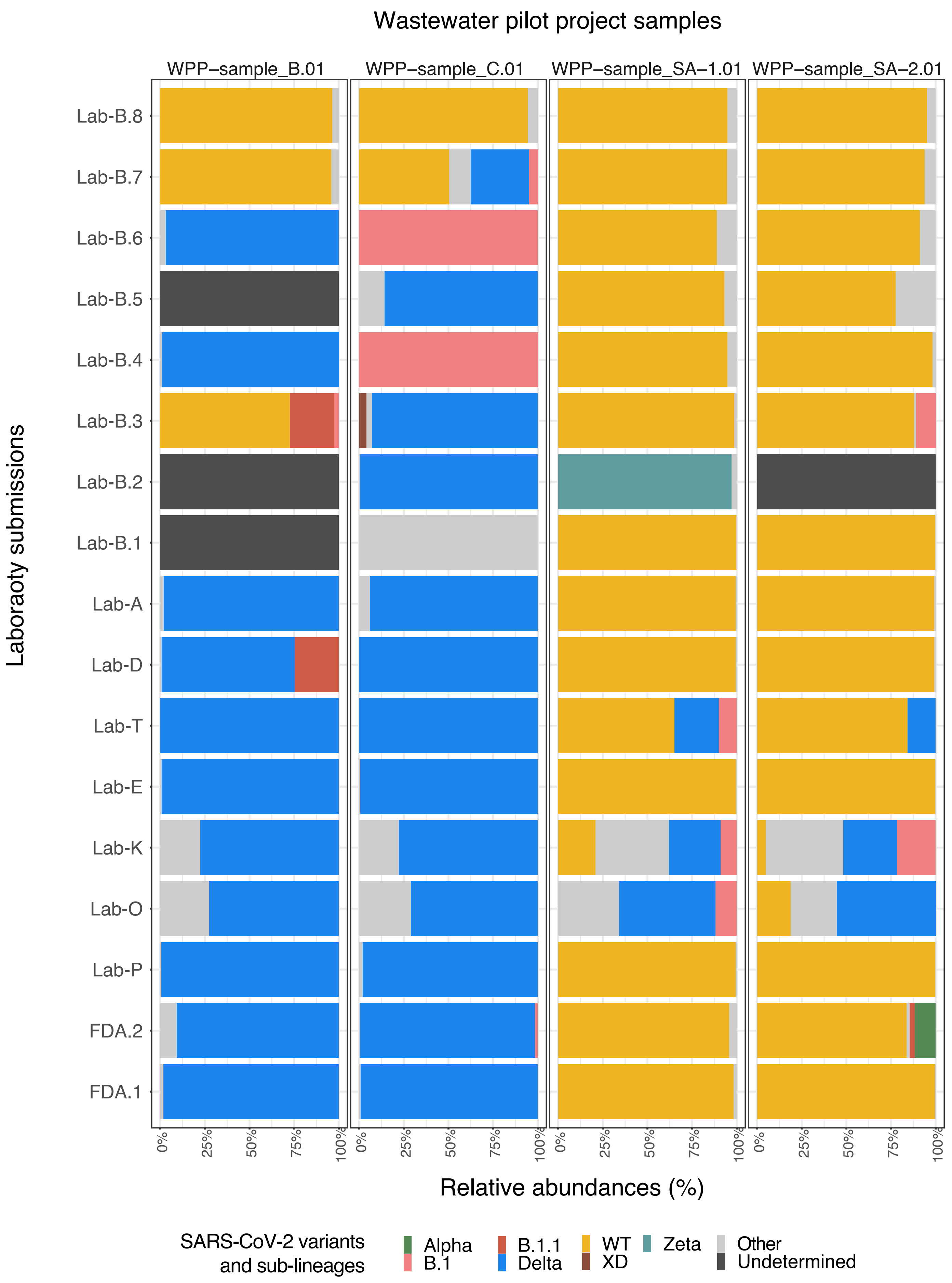

### Supplemental Figure 2

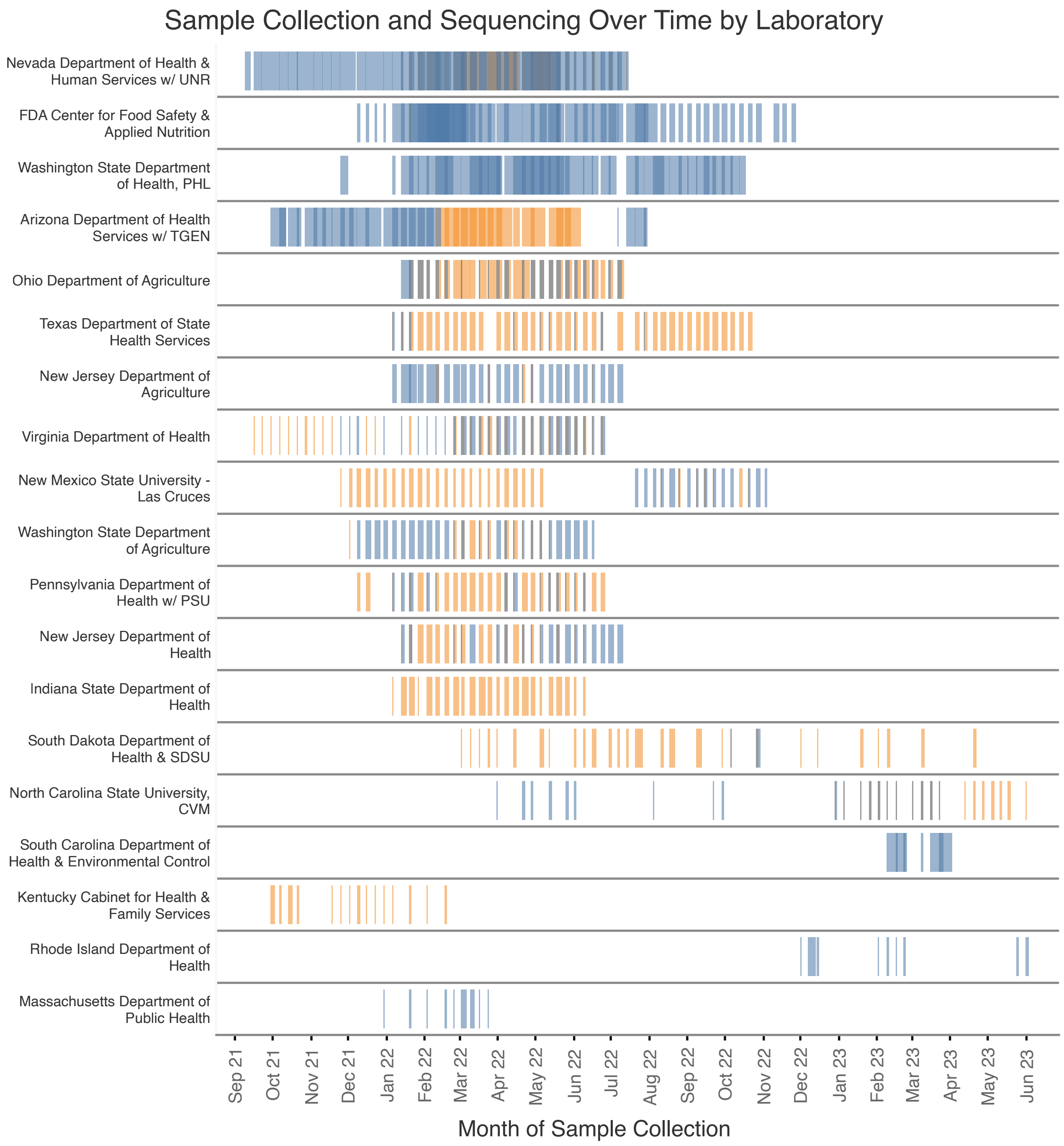
